# Neuronal-specific methylome and hydroxymethylome analysis reveal replicated and novel loci associated with alcohol use disorder

**DOI:** 10.1101/2023.11.28.23299094

**Authors:** Diego E. Andrade-Brito, Diana L. Núñez-Ríos, José Jaime Martínez-Magaña, Sheila T. Nagamatsu, Gregory Rompala, Lea Zillich, Stephanie H. Witt, Shaunna L. Clark, Maria C. Latig, Traumatic Stress Brain Research Group, PGC SUD Epigenetics Working Group, Janitza L. Montalvo-Ortiz

## Abstract

Alcohol use disorder (AUD) is a complex condition associated with adverse health consequences that affect millions of individuals worldwide. Epigenetic modifications, including DNA methylation (5mC), have been associated with AUD and other alcohol-related traits. Epigenome-wide association studies (EWAS) have identified differentially methylated genes associated with AUD in human peripheral and brain tissue. More recently, epigenetic studies of AUD have also evaluated DNA hydroxymethylation (5hmC) in the human brain. However, most of the epigenetic work in postmortem brain tissue has examined bulk tissue. In this study, we investigated neuronal-specific 5mC and 5hmC alterations at CpG sites associated with AUD in the human orbitofrontal cortex (OFC). Neuronal nuclei from the OFC were evaluated in 34 human postmortem brain samples (10 AUD, 24 non-AUD). Reduced representation oxidative bisulfite sequencing was used to assess 5mC and 5hmC at the genome-wide level. Differential 5mC and 5hmC were evaluated using the methylKit R package and significance was set at false discovery rate <0.05 and differential methylation >2. Functional enrichment analyses were performed and replication was evaluated replication in an independent dataset that assessed 5mC and 5hmC of AUD in bulk cortical tissue. We identified 417 5mC and 363 5hmC genome-wide significant differential CpG sites associated with AUD, with 59% in gene promoters. We also identified genes previously implicated in alcohol consumption, such as *SYK*, *CHRM2*, *DNMT3A*, and *GATA4*, for 5mC and *GATA4*, and *GAD1*, *GATA4*, *DLX1* for 5hmC. Replication was observed for 28 CpG sites from a previous AUD 5mC and 5hmC study, including *FOXP1*. Lastly, GWAS enrichment analysis showed an association with AUD for differential 5mC genes. This study reveals neuronal-specific methylome and hydroxymethylome dysregulation associated with AUD. We replicated previous findings and identified novel associations with AUD for both 5mC and 5hmC marks within the OFC. Our findings provide new insights into the epigenomic dysregulation of AUD in the human brain.

## 2 Introduction

The detrimental effects of alcohol use disorder (AUD) are substantial, resulting in more than 150,000 deaths globally (GBD 2019 Risk Factors Collaborators, 2020). AUD is characterized by persistent, uncontrollable, and excessive alcohol consumption despite its negative consequences. Although genome-wide association studies (GWAS) have identified genetic risk factors of AUD (Gelernter et al., 2019; Zhou et al. 2020, 2021, 2023), these only account for a portion of the variation observed.

Epigenetic mechanisms, such as DNA methylation (5mC), have been implicated in AUD and alcohol-related traits in human studies evaluating various tissues, including saliva, blood, and brain (Longley et al., 2021; Clark et al., 2022; Montalvo-Ortiz et al., 2022; Zillich et al., 2022). The 5mC mechanism involves the addition of a methyl group to the carbon 5 position of the nucleotide, which is catalyzed by DNA methyltransferases (DNMTs) (Gibney and Nolan, 2010). DNA hydroxymethylation (5hmC) occurs when this methyl group is removed through oxidation catalyzed by a family of ten-eleven translocase proteins (TET1, TET2, and TET3) during the DNA demethylation process. Recent work from our group and others has shown that 5hmC is functionally distinct from 5mC. This epigenetic mark is associated with transcriptional activation and highly prevalent in the brain (Rompala et al., 2022). Several studies have implicated 5hmC in anxiety-related behaviors (PMID: 28128679), schizophrenia, bipolar disorder (PMID: 25410542, PMID: 27411884), autism (PMID: 26423458), and Alzheimer’s disease (PMID: 33910000). Interestingly, a recent study evaluating 5mC and 5hmC in bulk tissue from the human postmortem brain identified a role for 5hmC in AUD (Clark et al., 2022).

Epigenetic patterns, such as 5mC and 5hmC, are tissue- and cell-type specific, and particularly 5hmC is highly enriched in the brain and abundant in neuronal cells, underscoring the need to investigate this epigenetic mark in brain tissue, particularly in neurons (Kriaucionis and Heintz, 2009; Szulwach et al., 2011; Mellén et al., 2012). However, most epigenetic studies have used bulk brain tissue, which can mask cell-type specific biological signals, highlighting the need for a cell-type-specific approach when evaluating the epigenetic landscape of AUD in the human brain.

The orbitofrontal cortex (OFC) has been implicated in decision making and motivated reward-related behavior (Morisot et al., 2019; Lohoff et al., 2021), and recent neuroimaging studies have associated alterations in this brain region with AUD (Shields and Gremel, 2020; Bracht et al., 2021; Atmaca et al., 2023). Individuals diagnosed with AUD exhibit a reduction in the OFC volume, accompanied by a decrease in gray matter, and an impact on dopaminergic pathways (Volkow et al., 2007, 2007; Coleman et al., 2011; Le Berre et al., 2014; Nimitvilai et al., 2017; Moorman, 2018; Morisot et al., 2019; Hernandez and Moorman, 2020). Recent 5mC studies from our group and others have revealed a role of epigenetic mechanisms in OFC in the context of substance use disorders (SUDs) (Kozlenkov et al., 2017; Rompala et al., 2022).

In this study, we examined neuronal-specific 5mC and 5hmC profiles in the OFC of AUD (n = 10) and non-AUD (n = 24) groups to identify epigenetically dysregulated genes and evaluate the differences between 5mC and 5hmC marks in the OFC. We also identify the functional pathways enriched by these epigenetically dysregulated genes, evaluate replication in an independent dataset, and assess its relationship with GWAS studies.

## 3 Materials and Methods

### 3.1 Sample collection

Our study cohort comprised 34 postmortem brain samples obtained from the National Post-Traumatic Stress Disorder (PTSD) Brain Bank51 (NPBB) (Friedman et al., 2017), a brain tissue repository at the U.S. Department of Veterans Affairs (VA). Consisting of European American and African American men with a mean age of 41 (s.d ± 12) (Friedman et al., 2017). The tissue samples were collected after obtaining informed consent from the next-of-kin and processed as described by (Friedman et al., 2017). The clinical diagnosis followed the antemortem assessment protocol (AAP) and postmortem diagnostic assessment protocol (PAP) based on the DSM-IV criteria (Friedman et al., 2017). The samples were categorized into AUD and non-AUD groups. The AUD group included 10 donors with alcohol use disorder (AUD) history, which refers to those diagnosed with alcohol dependence or alcohol abuse. The non-AUD group included 24 donors without an AUD diagnosis. AUD and non AUD groups were matched by posttraumatic stress disorder (PTSD), opioid use disorder (OUD), and current smoking. **Table 1** presents the demographic and clinical characteristics of the study cohort.

**Table 1.**
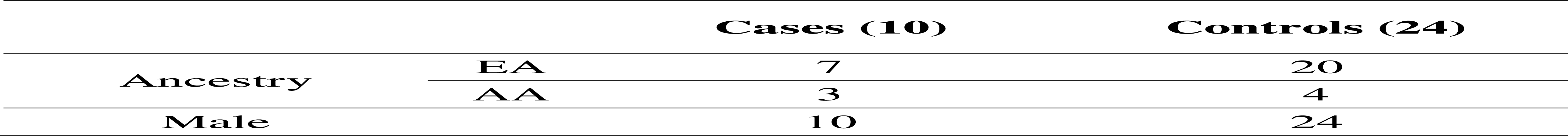
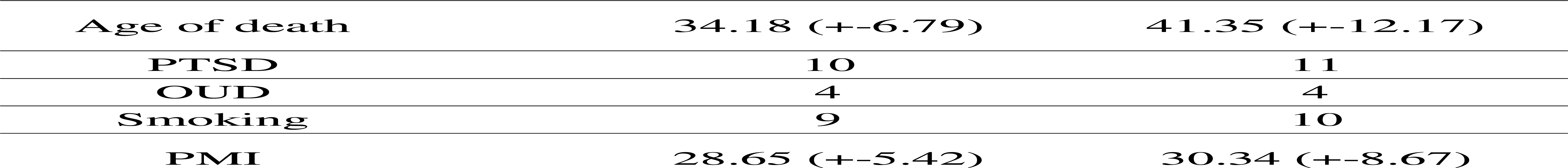
Demographic and clinical information of the study cohort.

### 3.2 Neuronal nuclei isolation and DNA extraction

Neuronal nuclei isolation was performed using fluorescence-activated nuclei sorting (FANS), described by Rompala and Nagamatsu et al. (2022) (Rompala et al., 2022), obtaining 0.5-1 M NeuN+ nuclei for DNA extraction. Sorted nuclei were centrifuged at 1500 × g for 15 min at 4°C to obtain a pellet. Next, 500 µL and 50 µL proteinase K (Cat. #69504, Qiagen, Valencia, CA) and 20 mg/mL RNAse A (Cat. #12091021; Thermo-Fischer, Waltham, MA) were used to refloat the pellet. TheDNeasy Blood and Tissue Kit (Cat. #69504, Qiagen) manufacturer’s protocol was used to process the samples. Finally, eluted samples were concentrated to a final volume of 20 µl with the Zymo Genomic DNA Clean and Concentrator-10 kit (Cat. #D4010, Zymo Inc., Irving CA) and stored at −80 °C.

### 3.3 High-throughput bisulfite sequencing and data processing

Sequencing data were obtained by reduced representation oxidative bisulfite-sequencing (RRoxBS), carried out at the Weill Cornell Epigenomics Core (New York, NY). The library preparations for 5mC and 5hmC were made using Mspl digestion for 400 ng of gDNA with the Ovation RRoxBS Methyl-Seq library preparation kit (TrueMethyl oxBS; Tecan, Switzerland). Bisulfite conversion was followed by a single-end 1×50 bp sequencing with the Illumina NovaSeq6000 system (mean depth of 42.7+/− 1.5 (µ +/− SEM) million reads per library). The hg38 genome reference was used for adapter trimming, alignment information, and mapping efficiency of the sequencing data using an in-house BiSeq pipeline. (Garrett-Bakelman et al., 2015).

### 3.4 Differential Methylation Analysis

The MethylKit R package (Akalin et al., 2012) was used to conduct differential methylation (5mC) and hydroxymethylation (5hmC) analyses at CpGs. The samples were filtered using a read coverage above 10x and under the 99.9^th^ percentile. Normalization was performed using the median method, where the mean was used to calculate the scaling factor to reduce coverage bias in the statistical analysis. Differential 5mC and differential 5hmC analyses were performed using logistic regression with correction for overdispersion and chi-squared significance testing (Akalin et al., 2012).The covariates included in the model were self-reported ancestry, age of death, PTSD, OUD, smoking, and postmortem interval (PMI). The sliding linear model (SLIM) was used to fit the p-values to q-values (Wang et al., 2011). The significance level for differential 5mC and 5hmC was defined as q-value < 0.05 and a greater than 2% difference of 5mC and 5hmC between AUD and non-AUD groups. Multiple correction sites (MTC) were used for annotation. We used the Genomation R package (Akalin et al., 2015) to annotate CpGs in promoters, introns, exons, and intragenic regions as well as CpG island, CpG islands shores, CpG shelves, and open sea (Akalin et al., 2015). Gene name annotation was performed using Ensembldb (Rainer et al., 2019).

### 3.5 Functional enrichment analysis

Genomation R package was used to perform genomic feature annotation (Akalin et al., 2015). Functional enrichment analysis was made using the gene annotation obtained by scan_region.pl perl tool and the UCSC genome browser annotation databases. Genes with a distance greater than 1500 bp were not considered for the enrichment analysis (Kuhn, Haussler, & James Kent, 2013). To conduct functional enrichment analysis we used Metascape (Zhou et al., 2019), Enrichr (Kuleshov et al., 2016), and WebGestalt (Liao et al., 2019), which integrates databases such as NCA TS BioPlanet (Huang et al., 2019), Panther (Mi et al., 2013), Gene Ontology Consortium (Gene Ontology Consortium, 2015), and the Kyoto Encyclopedia of Genes and Genomes (KEGG) (Kanehisa and Goto, 2000). Protein–protein interaction (PPI) enrichment analysis was conducted using the MTC of the differential 5mC and 5hmC. The molecular code detection (MCODE) algorithm was used to cluster enrichment ontology terms to identify neighborhoods where proteins are densely connected in the following databases: STRING, BioGrid, and OmniPath (Türei et al., 2016; Oughtred et al., 2019; Szklarczyk et al., 2023)

### 3.6 GWAS enrichment analysis

The software Multi-marker Analysis of GenoMic Annotation (MAGMA) v1.10 (de Leeuw et al., 2015) was used to conduct gene-level association analysis for 5mC and 5hmC using the GWAS data of alcohol use disorder (AUD) (Zhou et al., 2020), problematic alcohol use (PAU) (Zhou et al., 2020), cannabis use disorder (CUD) (Johnson et al., 2020), opioid use disorder (OUD) (Polimanti et al., 2020), and posttraumatic stress disorder (PTSD) (Nievergelt et al., 2019). The analysis was based on genetic variants in the 1000 Genomes Project dataset available on the MAGMA website (g1000_eur.bim). Gene annotation for the analysis was performed using the MAGMA NCBI37.3.gene.loc file.

An gene-level overlap between Clark et al. (2022) (Clark et al., 2022) reported genes, and our significant findings was conducted. Fisher’s exact test was applied using the GeneOverlap 1.38.0 R package (Li Shen, 2017) to evaluate whether the gene-level overlap is statistically significant.

## 4 Results

### 4.1 AUD-associated 5mC and 5hmC differential CpG sites

For 5mC, we identified 417 CpG sites after multiple testing correction with a difference in the percentage of methylation between AUD and non-AUD groups higher than 2. Of these, 137 were hypomethylated and 280 were hypermethylated (**Figure 1**). For 5hmC, we identified 363 CpG sites after multiple testing correction, with 213 hypo- and 150 hyper-methylated CpG sites (**Figure 2**). **Table 2** and **Table 3** list the top MTC differential 5mC and 5hmC CpG sites. The 5mC MTCs annotation showed that 59% of them were located at the promoter region, 21% at the intragenic region, 14% at introns, and 6% at exons. For the 5hmC MTCs, 66% were located at the promoter region, 17% at the intragenic region, 11% at introns, and 6% at exons.

**Figure 1.**
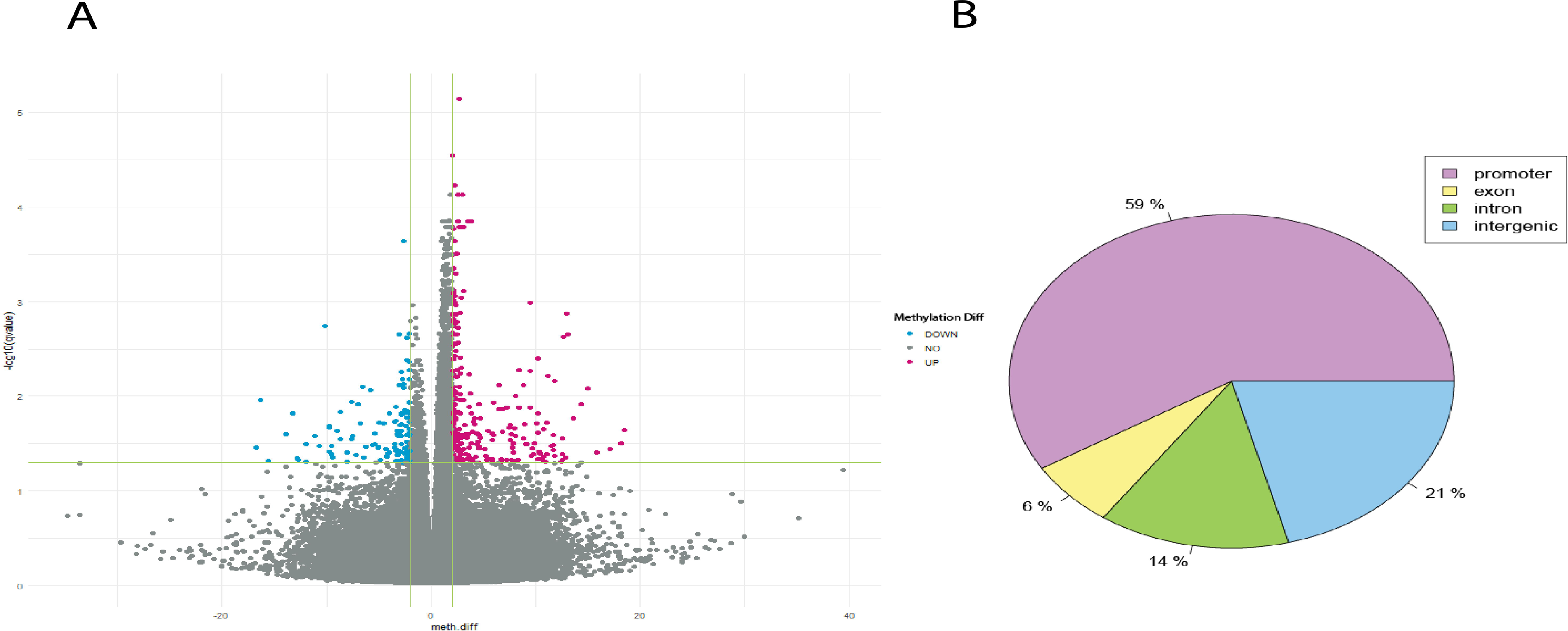
5mC differential CpG sites associated with AUD. A) Volcano plot shows the 5mC differential CpG sites associated with AUD. B) Pie chart depics the gene location of the MTC CpG sites identified.

**Figure 2.**
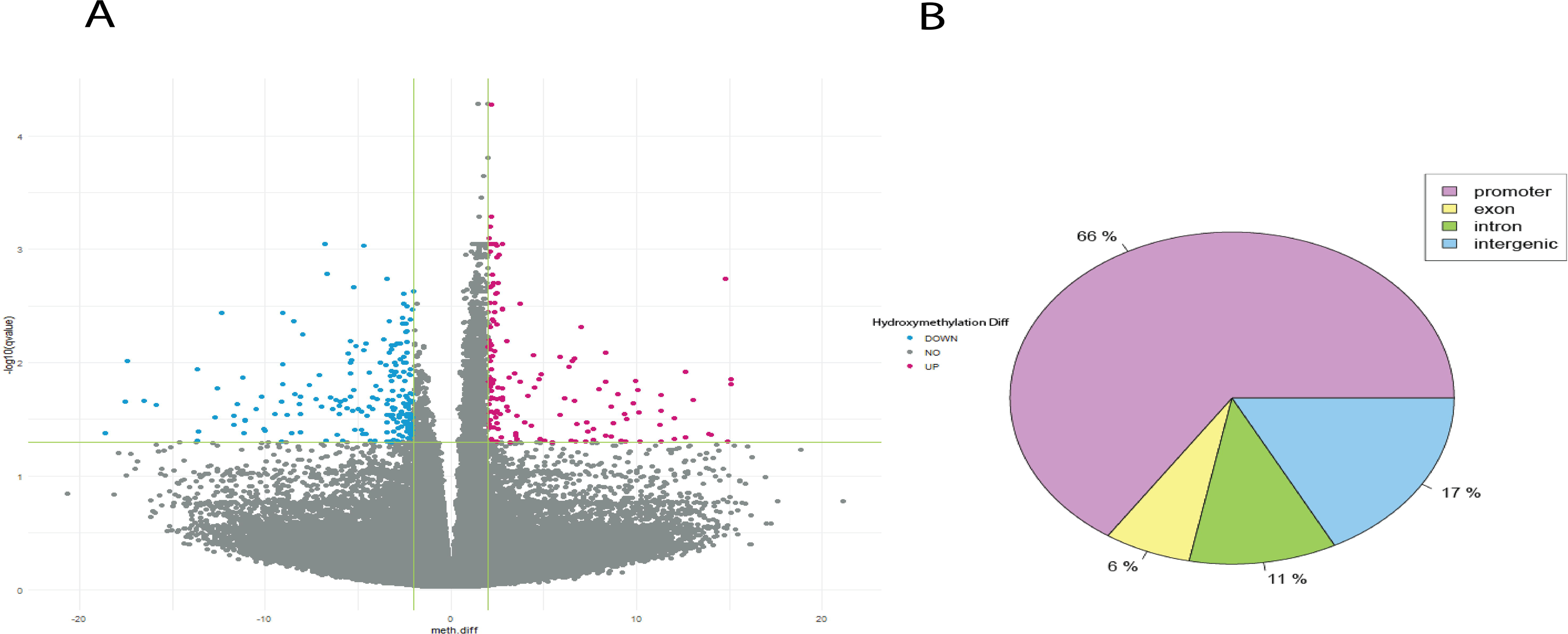
5hmC differential CpG sites associated with AUD. A) Volcano plot shows the 5hmC differential CpG sites associated with AUD. B) Pie chart depicts the gene location of the MTC CpG sites identified.

**Table 2.** Top MTC differential methylated (5mC) CpG sites.

| Gene | Chr | BP | strand | meth.diff | pvalue | qvalue |
| --- | --- | --- | --- | --- | --- | --- |
| *GNPNAT1* | chr14 | 52791636 | - | 2.73 | 3.93E-12 | 7.12E-06 |
| *CYP26B1* | chr2 | 72147339 | + | 2.02 | 3.17E-11 | 2.87E-05 |
| *PCDH8P1* | chr13 | 53201122 | - | 2.21 | 9.86E-11 | 5.95E-05 |
| *SNORA57* | chr11 | 62665457 | + | 3.04 | 2.46E-10 | 7.42E-05 |
| *FAM163B* | chr9 | 1.34E+08 | + | 2.55 | 2.03E-10 | 7.42E-05 |
| *KEAP1* | chr19 | 10503028 | - | 3.88 | 1.33E-09 | 0.000142 |
| *STAM2* | chr2 | 1.52E+08 | + | 2.55 | 1.20E-09 | 0.000142 |
| *ARX* | chrX | 25013092 | - | 3.48 | 1.22E-09 | 0.000142 |
| *AL033528.1* | chr1 | 25922969 | - | 2.70 | 1.76E-09 | 0.000162 |

**Table 3.** Top MTC differential hydroxymethylated (5hmC) CpG sites.

| Gene | chr | BP | strand | meth.diff | pvalue | qvalue |
| --- | --- | --- | --- | --- | --- | --- |
| *SLC12A8* | chr3 | 125141732 | + | 5.48 | 0.000104 | 0.049906 |
| *TFAP2A* | chr6 | 10419244 | + | -3.16 | 0.000104 | 0.049906 |
| *TLX1* | chr10 | 101135137 | - | -4.18 | 0.000103 | 0.049903 |
| *SOX17* | chr8 | 54453592 | + | -3.17 | 0.000103 | 0.049903 |
| *MAFA* | chr8 | 143431381 | + | 3.19 | 0.000103 | 0.049903 |
| *ZIC2* | chr13 | 99989614 | - | 11.33 | 0.000102 | 0.049745 |
| *RP11-570J4.2* | chr21 | 9592720 | - | 6.70 | 0.000102 | 0.04968 |
| *ARL6IP4* | chr12 | 122981331 | - | -3.29 | 0.000101 | 0.049664 |
| *PHOX2A* | chr11 | 72244124 | + | -3.20 | 0.0001 | 0.049519 |
| *NID2* | chr14 | 52068116 | + | -3.41 | 0.0001 | 0.049519 |

### 4.2 Q-Q plot

The quantile– quantile (QQ) plots (**Supplementary Figure 1**) for the 5mC and 5hmC differential analyses are shown. The lambda values were λ=1.21 for 5mC and λ=1.46 for 5hmC.

### 4.3 5mC and 5hmC-enriched pathways

For 5mC, we found significant enrichment (using 0bp annotation) for 31 pathways after multiple testing correction (**Supplementary Figure 2**). Top-level gene ontology (GO) pathways enriched included cellular process (GO:0009987), developmental process (GO:0032502), regulation of biological process (GO:0050789). The most significantly enriched pathways were cell-cell adhesion (GO:0098609), homophilic cell adhesion via plasma membrane adhesion molecules (GO:0007156), cell-cell adhesion via plasma-membrane adhesion molecules (GO:0098742), positive regulation of nervous system development (GO:0051962), regionalization (GO:0003002), and anterior/posterior pattern specification (GO:0009952). The enrichment of 5mC using the 1500bp annotation included 67 pathways (**Supplementary Figure 3**), with top-level GO pathways including cellular process (GO:0009987), developmental process (GO:0032502), and biological regulation (GO:0065007). The most significant enriched pathways were cell-cell adhesion (GO:0098609), homophilic cell adhesion via plasma membrane adhesion molecules (GO:0007156), cell-cell adhesion via plasma-membrane adhesion molecules (GO:0098742), anterior/posterior pattern specification (GO:0009952), regionalization (GO:0003002), pattern specification process (GO:0007389), and regulation of nervous system development (GO:0051960).

For 5hmC, significant enrichment using the 0bp annotation was found for 260 pathways (**Supplementary Figure 4**), including the top-level GO pathways of cellular process (GO:0009987), developmental process (GO:0032502), regulation of biological process (GO:0050789). After multiple testing correction, the most significantly enriched pathways were homophilic cell adhesion via plasma membrane adhesion molecules (GO:0007156), cell-cell adhesion via plasma-membrane adhesion molecules (GO:0098742), pattern specification process (GO:0007389), and cell-cell adhesion (GO:0098609). The enrichment analysis using the 1500bp annotation identified 165 significant pathways (**Supplementary Figure 5**), including top-level GO pathways enriched for 5hmC of cellular process (GO:0009987), developmental process (GO:0032502), growth (GO:0040007). The most significant enriched pathways were homophilic cell adhesion via plasma membrane adhesion molecules (GO:0007156), cell-cell adhesion via plasma-membrane adhesion molecules (GO:0098742), pattern specification process (GO:0007389), regionalization (GO:0003002), and cell-cell adhesion (GO:0098609).

### 4.4 Protein-protein interaction analysis

The PPI network analysis of the differential 5mC genes showed as significant pathways cell-cell adhesion (GO:0098609), homophilic cell adhesion via plasma membrane adhesion molecules (GO:0007156), cell-cell adhesion via plasma-membrane adhesion molecules (GO:0098742). The MCODE cluster algorithm identified GO pathways related to neurogenesis, chromatin organization, and cell adhesion (**Supplementary Figure 6** and **Supplementary Figure 7**).

For the 5hmC marks, the significant pathways identified in the PPI network analysis were cell-cell adhesion homophilic cell adhesion via plasma membrane adhesion molecules (GO:0007156), embryonic organ development (GO:0048568), and cell–cell adhesion via plasma-membrane adhesion molecules (GO:0098742). The MCODE cluster algorithm identified GO pathways implicated in neurogenesis, cell adhesion, calcium ion transport, and Wnt signaling (**Supplementary Figure 8** and **Supplementary Figure 9**).

### 4.5 GWAS enrichment results

A significant enrichment was identified between genes with differential 5mC and GWAS signals of AUD (p= 0.0022) and PAU (p = 0.019). No significant enrichment of alcohol-related GWAS was observed for 5hmC or for CUD, OUD, or PTSD with either 5mC or 5hmC (**Figure 3**).

**Figure 3.**
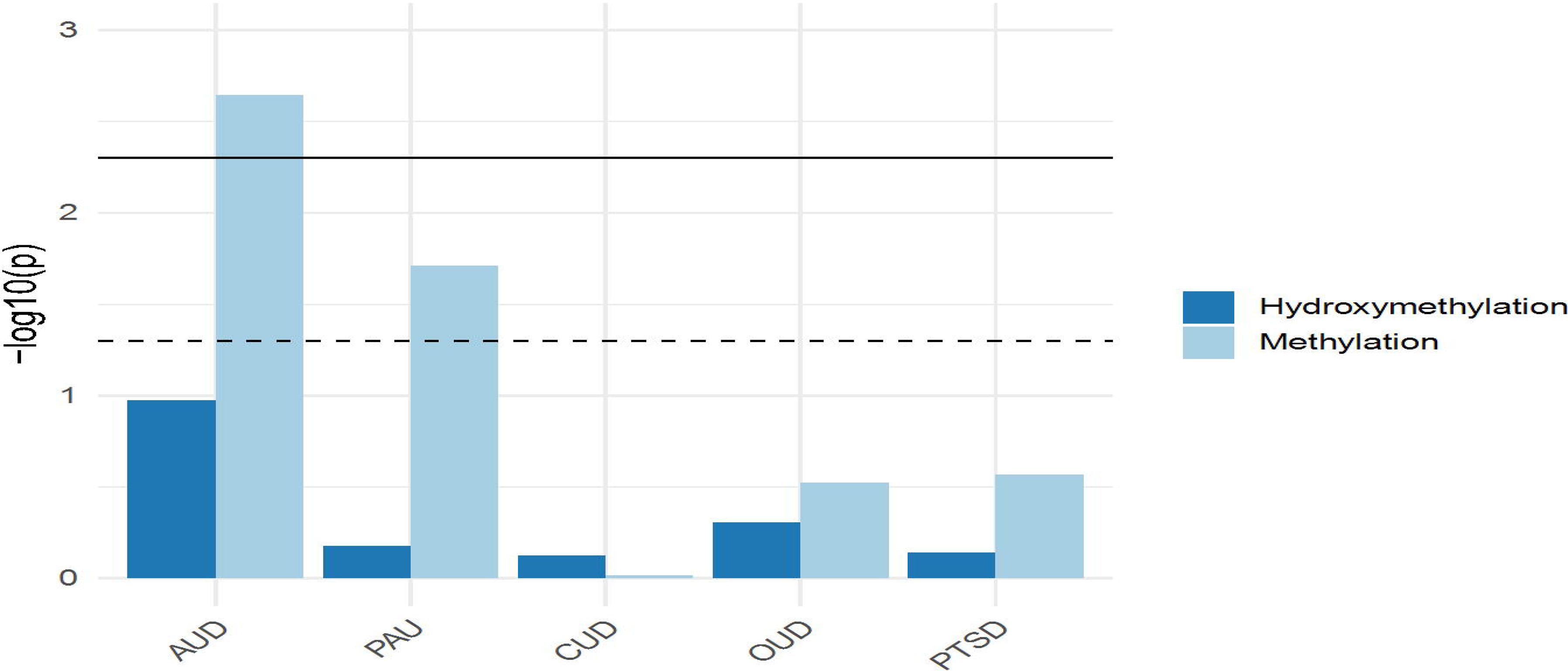
Generalized Gene-Set Analysis of GWAS for 5mC and 5hmC. The bar plot shows the gene-set analysis of GWAS for 5mC and 5hmC, including alcohol use disorder (AUD), problematic alcohol use (PAU), cannabis use disorder (CUD), opioid use disorder (OUD), and post-traumatic stress disorder (PTSD).

### 4.6 Gene overlap analysis

We compared our findings with those reported by Clark et al. (2022) (Clark et al., 2022), which evaluated AUD-associated 5mC and 5hmC in bulk tissue from the human postmortem PFC. For 5mC, we found an overlap of 14 genes with their reported 576 (Figure 4A). Fisher exact test showed that the overlap is significant (p = 8.5e-25, odds ratio=124.00). Similarly, for 5hmC, we found a significant overlap of 14 genes with their reported 1023 genes (Figure 5B; p = 3.9e-22, odds ratio=79.80).

**Figure 4.**
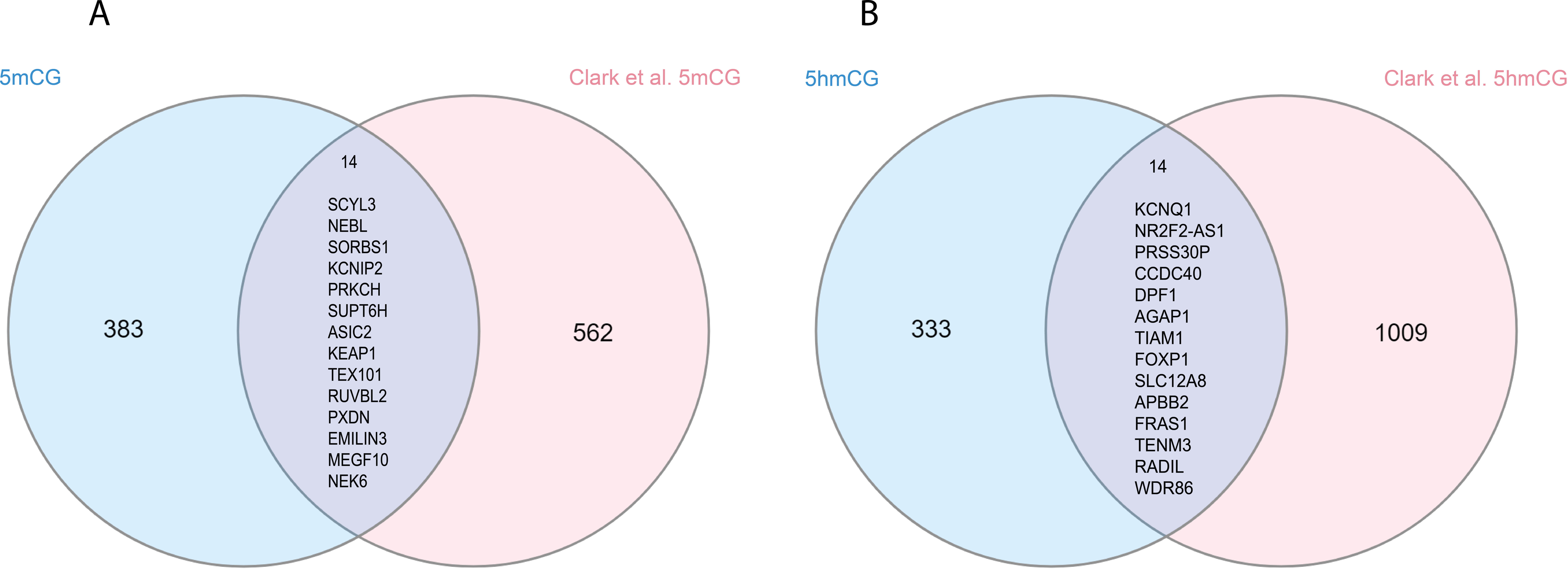
Overlapped differential methylated (5mC) and hydroxymethylated genes in human brain tissue. A) shows the overlap between our 5mCG MTCs and 5mCG reported sites from Clark et al. (2022), where 14 genes overlap between both results. B) shows the overlap between our 5hmCG from OFC and 5hmCG reported sites from Claret al. (2022), where 14 genes overlap between both results.

## 5 Discussion

This study presents a neuronal-specific epigenomic investigation of AUD in the human brain. We profiled 5mC and 5hmC at the genome-wide scale and revealed previously reported as well as novel differential CpG sites associated with AUD. Our analysis identified 417 and 363 AUD-associated CpG sites for 5mC and 5hmC, respectively.

Our 5mC findings revealed several genes of interest in the AUD context, including *SYK*. This gene has been previously associated with alcohol metabolism in the liver and its inhibition is linked to reduced liver inflammation (Qu et al., 2018; Kurniawan et al., 2020). Furthermore, previous studies have found that binge drinking can induce SYK activation, and pharmacological inhibition of SYK significantly decrease alcoholic liver disease in a mouse model of binge drinking (Bukong et al., 2017).

Another differential 5mC gene identified is the *DNMT3A*, which encodes for an enzyme involved in de novo DNA methylation (Fischer et al., 2021; Pino et al., 2017). Ethanol exposure can induce a prolongued upregulation of *DNMT3A* in neuronal precursor cell lines and primary mouse embryonic fibroblasts (Miozzo et al., 2018). Similarly, *Dnmt3a* has been found upregulated in the nucleus accumbens of alcohol-preferring rats exposed to intermittent ethanol exposure (Niinep et al., 2021).

For 5hmC, one of the identified differential CpG sites previously linked to AUD mapped to *GAD1*, a gene that encodes for one of the glutamate decarboxylases that catalyze the conversion of glutamate to Gamma-aminobutyric acid (GABA). Chronic alcohol exposure has been linked to a decrease in GABA levels and an increase in GABA receptors (Sytinsky et al., 1975; Tran et al., 1981; Dodd et al., 1996; Behar et al., 1999). *GAD1* has also been found to be upregulated in the dorsomedial thalamus of human subjects in individuals with AUD (Hade et al., 2021). However, a recent study did not find a difference in *GAD1* mRNA levels in OFC between individuals with AUD and the non-alcoholic group AUD (Underwood et al., 2019; Edenberg et al., 2010). More research is needed to fully elucidate the role of these genes in AUD.

Additional AUD-associated CpG sites with differential 5hmC were those mapped to *DLX1* and *DLX2*, which are also implicated GABA signaling, specifically in interneuron GABA synthesis (Le et al., 2017; Pla et al., 2018) and play a role in interneuron synaptogenesis and dendritogenesis (Pla et al., 2018). Moreover, these genes are suggested to directly promote the expression of *Grin2b*, a gene previously reported in human steam-cell-derived cortical neurons exposed to chronic alcohol consumption and also reported in the prefrontal cortex and hippocampus of mice treated with chronic alcohol consumption followed by withdrawal (Endele et al., 2010; Xiang et al., 2015; Pla et al., 2018; Myers et al., 2019). In the current study, both *DLX1* and *DLX2* showed hyper-5hmC and were located in the exon region. *DLX2* CpG site was located in the promoter region, suggesting that it could be directly impacting gene expression regulation in individuals with AUD. The effect of 5hmC in the exon region is not well understood; however, in the case of 5mC, it is suggested that the density of 5mC in the exon can enhance gene expression (Li et al., 2018).

*GATA4* was identified in both 5mC and 5hmC differential analyses and has been previously associated with alcohol dependence (Treutlein et al., 2009; Edenberg et al., 2010; Karpyak et al., 2014). *GATA4* encodes the GATA-motif binding protein type 4, a transcription factor that controls the expression of proteins involved in drug metabolism (Karpyak et al., 2014). In addition, high doses of alcohol increase its expression (Zhong et al., 2010). In our study, this gene was hypomethylated and hypo hydroxymethylated (5mC location at 11697675bp, 5hmC location at 11703255). The differential 5mC CpG site was located in the intronic region, suggesting chromosomal instability. The 5hmC CpG sites were located in the promoter and intronic regions. The presence of 5hmC at the promoter region has been associated with protection of gene transcription in regions where 5mC is present (Ehrlich and Ehrlich, 2014). This suggests an interaction of these epigenetic mechanisms in the modulation of *GATA4* expression in the OFC.

The findings of our enrichment analysis are consistent with prior AUD-related studies identifying development, and neurogenesis. For instance, in a previous report examining fetal alcohol syndrome (FAS), these pathways were found to be highly significant (Fischer et al., 2021). Moreover, studies have also reported that AUD impacts cell adhesion and neurogenesis, which involves the development of new neurons and their integration into functional neural networks (Ramanathan, 1996; Arevalo et al., 2008; Pino et al., 2017; Poulose et al., 2017; Lees et al., 2020; Wooden et al., 2021). The effects on developmental processes and neurogenesis may contribute to the cognitive impairment reported in individuals with AUD and may also be linked to brain dysfunction in various regions, including the OFC (Arevalo et al., 2008; De Wilde et al., 2007).

GWAS enrichment analysis showed enrichment of differential 5mC genes with AUD and PAU GWASes (Zhou et al., 2020). No enrichment was identified with GWAS of other SUDs, suggesting a specificity of our differential 5mC marks with alcohol-related traits. When comparing our findings with those reported by Clark et al. (2022), we observed 14 overlapping genes for each epigenetic mark, 5mC and 5hmC. Several overlapping genes with differential 5hmC have been previously linked to SUDs. For instance, *KCNQ1,* a gene encoding a potassium ion channel, was identified in a genome-wide association study (GWAS) of alcohol dependence (Edenberg et al., 2010; Feng et al., 2022). Similarly, *APBB2* has been associated with opioid and amphetamine dependence (Gelernter et al., 2014; Liu et al., 2018). Among the 5mC overlapping genes, *ASIC2* has been linked to addiction-related behavior in mice (Kreple et al., 2014). These findings underscore the importance of evaluating both 5mC and 5hmC to fully investigate the associations between AUD and epigenetic mechanisms, at least in the brain where 5hmC is highly prevalent and enriched in neurons. Future studies should explore these genes in greater depth to better understand their involvement in AUD. The limitations of this study are that the donors of the study cohort present heterogeneous comorbidities. All individuals in the OUD group were also diagnosed with PTSD. We controlled this by using PTSD as a covariate in the differential 5mC and 5hmC analyses. In addition, we conducted a GWAS enrichment analysis of our 5mC and 5hmC annotated genes to determine whether the reported genes were enriched for the comorbidity traits, including PTSD and other SUDs. In addition, the cohort size is limited; however, it is comparable to other recently published postmortem brain studies. Another limitation is that all samples are male, limiting to identify the effect of sex in our results. The analyses were only carried out on CpGs sites and it would be important to conduct a study on non-CpGs sites, because of the role of 5mC at non-CpGs on neuropsychiatric diseases from our group and others (Jang et al., 2017; Nagamatsu et al., 2022). To determine if the epeigenetic marks in the reported genes in this study are a cause of AUD or an effect, it is necessary to conduct research in model organisms.

Our study characterized the methylome and hydroxymethylome profiles of AUD in neurons from the OFC. Our results replicate previous findings in certain genes and highlight new findings for both 5mC and 5hmC. This study reveals new insights into the epigenomic dysregulation of AUD in the human brain and pinpoints potential drug targets for the treatment of individuals suffering from AUD.

## Data Availability

The original contributions presented in the study are included in the article/supplementary material,
further inquiries can be directed to the corresponding author/s

## 10 Acknowledgments

We want to thank the donors and their families for making this research possible.

**Members of the Traumatic Stress Brain Research Group:** Victor E. Alvarez, MD; David Benedek, MD; Alicia Che, PhD; Dianne A. Cruz, MS; David A. Davis, PhD Matthew J. Girgenti; PhD, Ellen Hoffman, MD, PhD; Paul E. Holtzheimer, MD; Bertrand R. Huber, MD, PhD; Alfred Kaye, MD, PhD; John H. Krystal, MD; Adam T. Labadorf, PhD; Terence M. Keane, PhD; Mark W. Logue, PhD; Ann McKee, MD; Brian Marx, PhD; Deborah Mash, MD; Mark W. Miller, PhD; Crystal Noller, PhD; Janitza Montalvo-Ortiz, PhD; William K. Scott, PhD; Paula Schnurr, PhD; Thor Stein, MD, PhD; Robert Ursano, MD; Douglas E. Williamson, PhD; Erika J. Wolf, PhD, Keith A. Young, PhD

Members of the SUD PGC

## 12 Supplementary Material

1. Differential CpG Methylation (FDR-adjusted q<0.05)
2. Differential CpG Hydroxymethylation (FDR-adjusted q<0.05)
3. GWAS enrichment analysis results
4. Metascape enrichment analysis results for Methylation
5. Metascape enrichment analysis results for Hydroxymethylation
6. Overlapped MTC genes vs Clark et al. (2022) for Methylation
7. Overlapped MTC genes vs Clark et al. (2022) for Hydroxymethylation

## 13. Data Availability Statement

Summary statistics are available in Supplementary Tables. Summary-level data produced in the present work are included in the Supplementary materials. All data produced are available upon reasonable request.

